# Executability and repeatability of a study setup using wearable and computerized technology to examine a wide range of physiological and cognitive functions of a patient outside hospital

**DOI:** 10.1101/2023.01.30.23285187

**Authors:** Kiti Müller, Ilari Rautalin, Lauri Ahonen, Anne Arola, Andreas Henelius, Hanna Jokinen, Jussi Korpela, Miikka Korja, Nicolas Martinez-Majander, Aaro Mustonen, Teemu Paajanen, Satu Pakarinen, Kati Pettersson, Jukka Putaala, Laura Sokka, Tuomas Tikka, Jussi Virkkala

**Author notes:** Corresponding author: Dr Kiti Müller.

## Abstract

**Objectives:** Reasons for a patient’s daily problems are often unclear as they are not systematically monitored in daily life. To remedy this, we designed a study protocol combining outpatient laboratory and home measurements. Feasibility of the protocol was evaluated by examining its executability and repeatability.

**Participants:** Sixteen patients undergoing neurosurgery for unruptured intracranial aneurysm (UIA), each acting as one’s own control.

**Study Protocol:** Two laboratory examinations followed by home recordings at 2 weeks pre- and 4-6 weeks postoperatively. In between a sole home recording at 3 weeks and 3 months postoperatively. Subjective health and performance data were gathered via an internet portal.

**Methods:** Laboratory Day included computer-based cognitive tests on attention, working memory and executive functions. Electroencephalography (EEG), electrocardiography (ECG) and electrodermal activity (EDA) were recorded during the tests. A saccade test with electro-oculography (EOG) and event-related potentials (ERP) to emotionally loaded sounds were also done. First three home recordings included tablet-based cognitive tests, sleep polysomnography (PSG), ECG, and actigraphy; the 4^th^ only an actigraphy and sleep diary.

**Outcome measures:** Executability and repeatability of the protocol: Number of patients completing the study, success rate of carrying it out as planned, obtained data and patient feedback.

**Results:** In 13 (81%) patients all data from home recordings was obtained as planned. In 11 (68%) patients the whole protocol was carried out as planned and in 3 partly. The time schedules required adjusting. Patients reported no discomfort from using wearable devices. Patients considered answering the internet-based health questionnaires cumbersome: all data was obtained only in 30%.

**Conclusions:** Mobile technologies are a tool kit from which to choose a clinically relevant combination to obtain objective data on a patient’s physical, physiologic, and cognitive performance outside hospital.

**Strengths:**

- The study consisted of a wide range of medical devices providing objective information on different physiologic functions and cognitive performance of a patient relevant for daily activities.
- The research examined executability, repeatability and patient compliance of a clinical study protocol and aspects relevant to obtaining clinically useful data.

**Limitations:**

- Participants did not have any chronic diseases that could affect especially home recording success
- The study setup and protocol were not tested outside the home environment.
- The ability of patients to set up the different measurements independently and correctly was not tested.

## INTRODUCTION

Even though clinical examination at a treating clinic shows favorable response to medical treatment, physical recovery after an operation or rehabilitation is assessed as good, patients often describe prolonged difficulties in daily activities. Results of medical examinations on a patient’s performance done at this point are usually compared to test norms due to lack of data on the patient’s own performance level prior to a medical intervention. Therefore, minor decrements of function, particularly in high-functioning individuals, may remain undetected. In addition, used methods seldom provide information on performance fluctuations as their duration is too short for this.

Information on how a patient is coping with everyday life and working activities is typically based on patient interviews and different types of quality-of-life [1] and working ability [2] questionnaires. Subjective evaluations do not always reflect actual human performance. Individuals describe and grade the severity and effects of symptoms on daily activities differently.

Currently different aspects of performance of patients are not systematically monitored outside hospital or outpatient centers. However, several mobile medical technologies are nowadays so mature that they can be safely used in everyday life. Data obtained with these devices provide objective information on a wide range of human performance aspects.

We designed a study protocol that combines hospital, outpatient laboratory and home measurements of cardiovascular and central nervous system physiology, cognitive functions, and sleep using mobile computerized and wearable devices. The protocol can be repeated, and performance data gathered during a patient’s follow-up can be compared to the earlier obtained results.

To test the executability and repeatability of the protocol we recruited patients undergoing neurosurgery for unruptured intracranial aneurysm (UIA). The operation is a clear intervention time point and allows pre- and postoperative examination using the same study protocol. Also, while the rate of neurosurgical complications in UIA [3] is low and even though physical recovery assessed at clinical control is good, patients frequently report reduced quality of life and experience difficulties in returning to work [4].

The executability and repeatability of the protocol was tested with otherwise healthy UIA patients. Of interest was also how patients commit to repeated measurements taking place during recovery from an operation

## METHODS

This study was carried out by the Department of Neurosurgery, Helsinki University Hospital (HUS) and the Finnish Institute of Occupational Health (FIOH). All patients provided written consent after being informed of the study. The study followed ethical principles of the Declaration of Helsinki and was approved by the Ethics Committee of Helsinki and Uusimaa Hospital District (38/13/03/00/2016).

### Study subjects

From 100 potentially eligible UIA patients, 16 patients, 3 men and 13 women, age range 26 – 61 yrs., mean 46 yrs., were enrolled. Inclusion criteria were: (1) radiologically (computed tomography with angiography) confirmed unruptured intracranial aneurysm(s) (UIA) with a maximum diameter of ≥ 3 mm, (2) listed on an elective surgical (not endovascular) waiting list by a cerebrovascular neurosurgeon, (3) age between 25 and 62 years, (4) working at the time of enrollment, (5) resident within 80 kilometers (50 miles) of the study hospital, and (6) Finnish speaking. Exclusion criteria were: 1) previous treatment of UIA, 2) ongoing psychiatric disorder, and 3) history of alcohol and/or substance abuse. Recruitment flow chart can be found in online <u>supplemental file 1</u>.

Of the recruited patients, 3 had experienced a brain transient ischemic attack (TIA) and 1 intracerebral bleeding. These neurologic events triggered their further neurologic examinations revealing the UIAs in their brain scans. All four had recovered well from the primary disease incident. One patient had earlier been treated for encephalitis in childhood and another for meningitis in 2015 and 2016. Both had recovered fully without any residual symptoms.

The eligible participants were first approached by phone and then visited a neurologist at the hospital to discuss the protocol in detail and go through the written information of the project.

They then gave written consent to voluntary participate. They were also informed that they were free to withdraw from the study at any time without any effect on the treatment of their UIA. Giving a reason for withdrawal was voluntary.

### Study protocol

All recruited UIA patients were operated according to the standard treatment protocol with one exception: Brain magnetic resonance imaging (MRI) was performed both pre- and postoperatively to detect possible surgery-related complications.

The study timeline is shown in figure 1. Examinations included pre- and postoperative clinical examinations at HUS, outpatient laboratory tests at FIOH, and home measurements. 1^st^ and 3^rd^ outside hospital examination days consisted of both laboratory examinations and home recordings and the 2^nd^ test day only home recordings. The 4^th^ measurement was a home recording with a reduced setup. The planned time points for the examinations out-of-hospital were as follows: 1^st^ at 2 weeks before the operation, 2^nd^ at 3 weeks after and 3^rd^ approximately 6 weeks after the operation and near expected end of sick leave and return to work. The 2^nd^ test daytime was chosen based on clinical experience of neurosurgeons: Postoperative recovery starts to slow down after 3 weeks in some patients. The 4^th^ data gathering was planned for 3 months postoperatively, near the time of clinical follow-up at HUS neurosurgery department.

**Figure 1.**
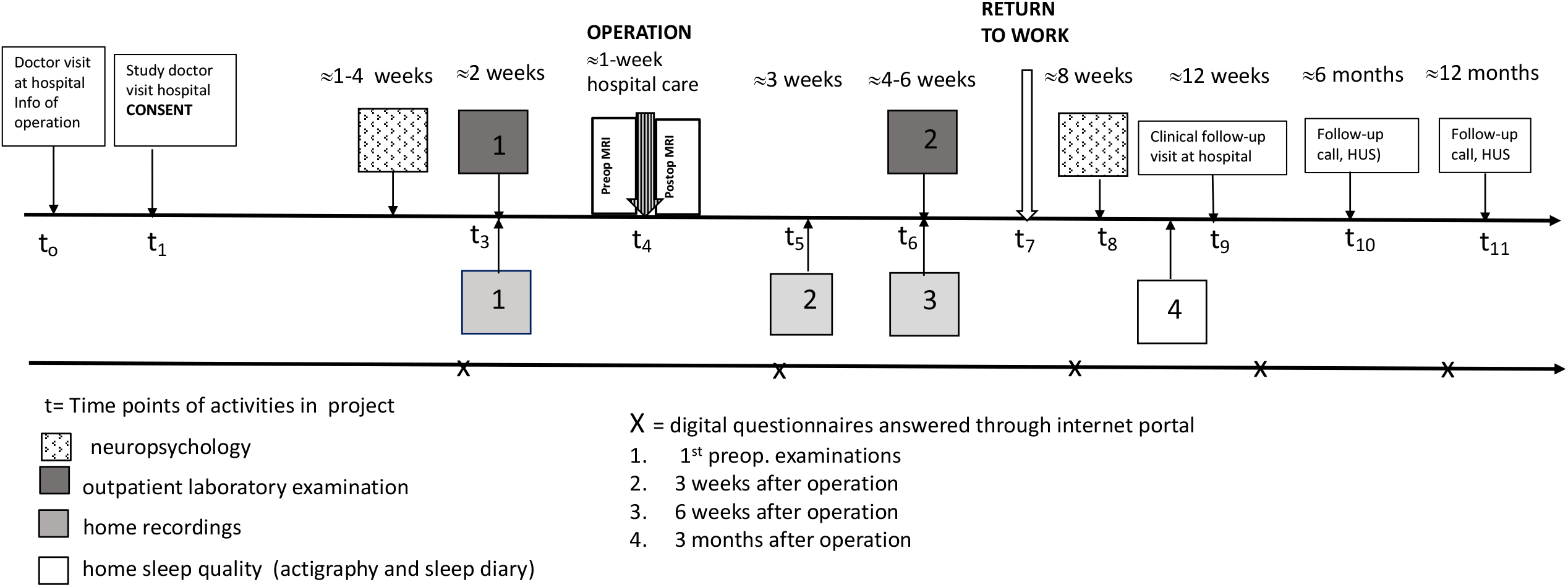
Overview of the study protocol showing the planned time-points for the different examinations in relation to surgery. Planned timeline of the study protocol.

Outside the actual methodological study protocol, research nurses contacted the patients by phone to fill in a questionnaire on their current health and work status at 6 months and again at 12 months when follow-up ended. In addition, patients filled subjective questionnaires (described in the online <u>supplemental file 3</u>) via an internet portal.

### Hospital examinations

#### Neuropsychological assessments

Patients underwent comprehensive neuropsychological examinations 1-4 weeks before and two months after surgery. The following domains were examined with standard paper and pencil tests: processing speed, executive functions, working memory, memory and learning, verbal functions, and visuo-perceptual abilities. In addition, a recently developed computer-based Flexible Attention Test (FAT) battery was included. The FAT subtests are modified from the classical neuropsychological tests Trail Making Test [5]and Corsi Block Tapping task [6]. Validated questionnaires were used to evaluate insight and awareness of cognitive symptoms, cognitive reserve, fatigue, psychological well-being, and resilience. More information and references for the used methods are presented in <u>online supplemental file 3</u>.

### Examinations outside hospital

#### Laboratory measurements

The content of the laboratory day is depicted in figure 2. Each day started with a questionnaire about patient’s past 24-hour activities, sleep, stress, medication, smoking, alcohol, and caffeine consumption and work.

**Figure 2.**
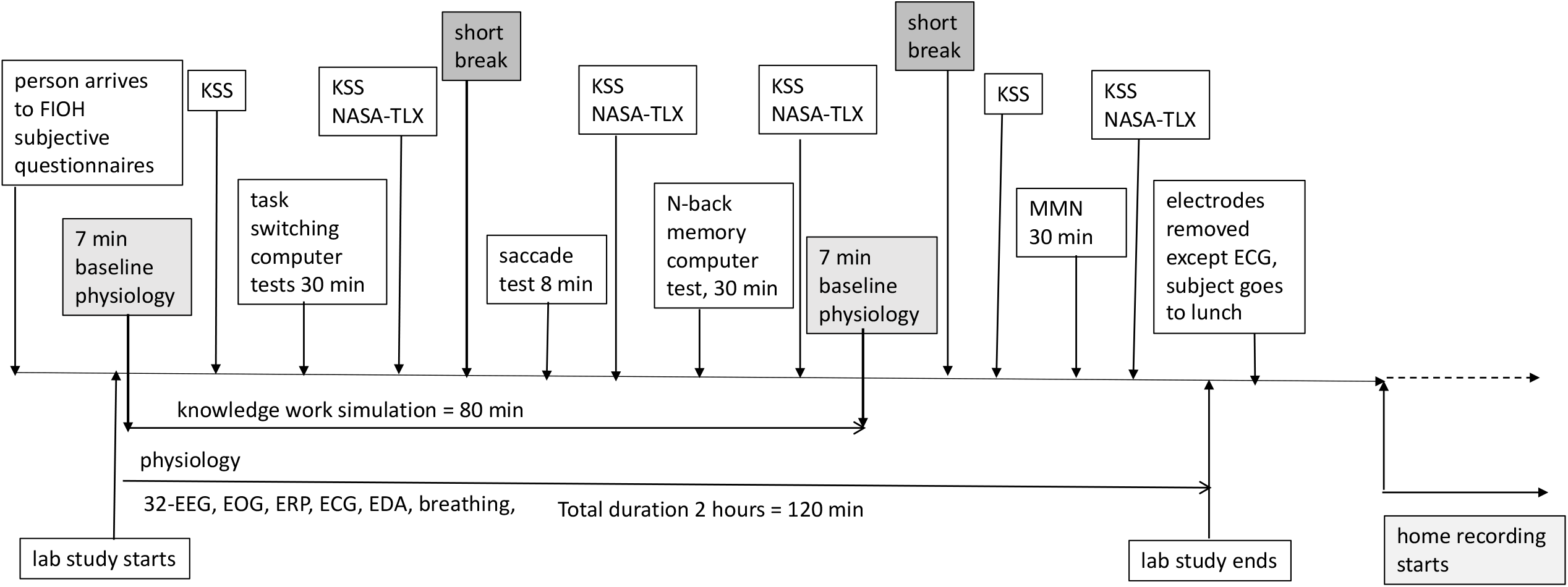
The laboratory study protocol. Laboratory study protocol at FIOH (Finnish Institute of Occupational Health). EEG = electroencephalography, EOG = electro-oculography, ERP = event-related potentials. ECG = electrocardiography, EDA = electrodermal activity, NASA-TLX = Nasa Task Load Index, KSS = Karolinska Sleepiness Scale, MMN = multifeatured mismatch negativity

The laboratory study session was designed to simulate an 80-minute working session in which participants were required to carry out computerized cognitive tasks with varying difficulty. The N-back [7] and Task-switching [8] tasks were applied as they are extensively used for measuring attention, working memory and cognitive flexibility/executive functions, which are crucial for carrying out work and everyday life tasks. Task difficulty was varied to examine the effects of workload on cognition.

The patient’s autonomic nervous system (ANS) and central nervous system (CNS) physiologic state affects cognitive performance and vice versa[9]. Electrocardiogram (ECG) and electrodermal activity (EDA) were recorded to obtain data on ANS and electroencephalography (EEG) on CNS while patients carried out the computer-based tasks. Scalp recordings of event-related potentials (ERP) were used to study brain cortical processing associated with the tasks. The start and end time of the task sessions were marked. The physiologic data was annotated to identify in which type of task the data was gathered. In addition, we measured eye movements with electro-oculography (EOG) during a saccade test [10]. The test provides information on frontal lobe guided attention. Eye movements also give information on vigilance and sleepiness [11]. Figure 3a shows the placement of physiologic sensors on subjects during laboratory tests.

**Figure 3.**
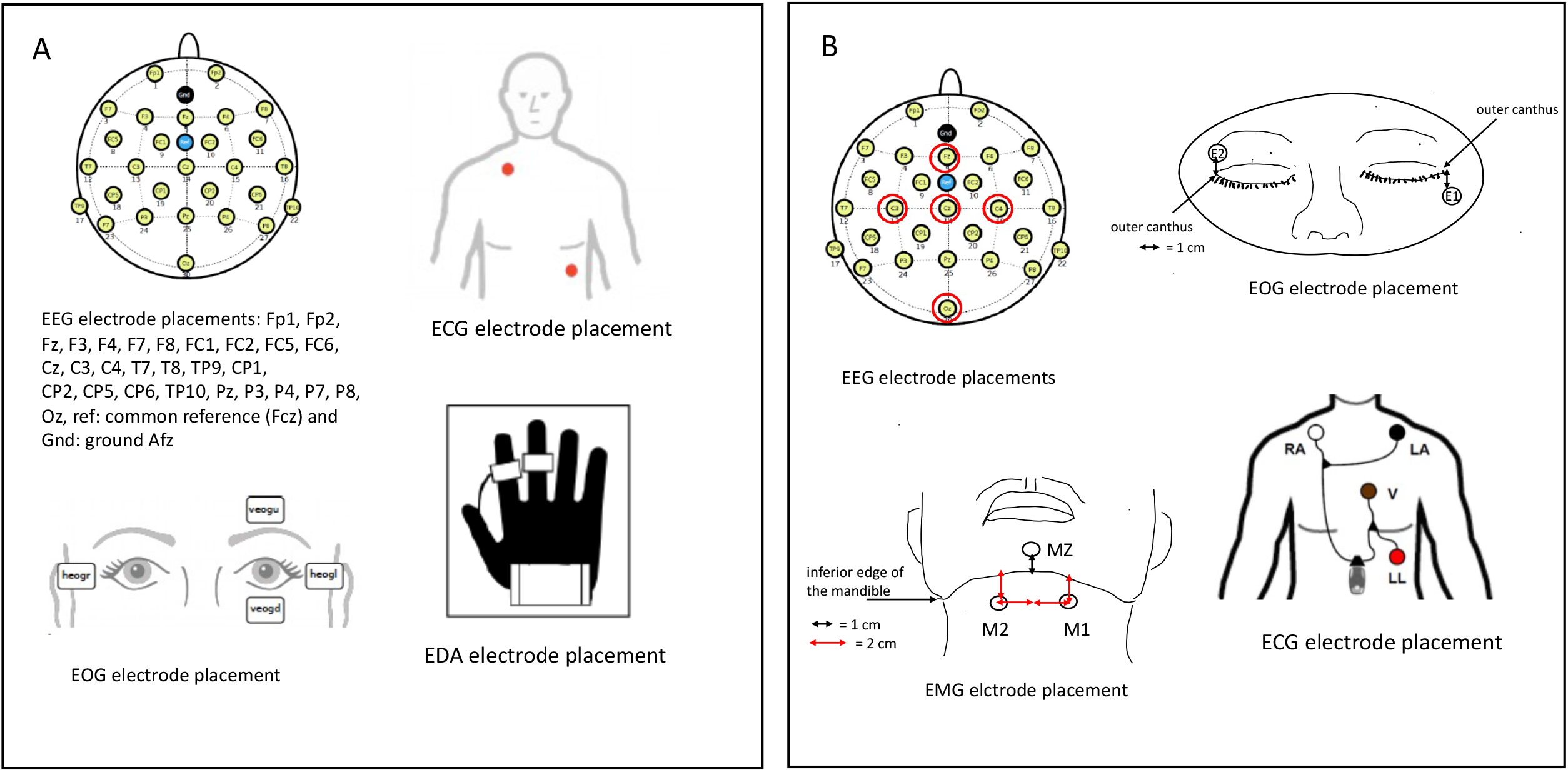
Electrode placements. The placements of physiologic sensors on the subject in the laboratory (A) and home (B) examinations: 32-channel electroencephalography (EEG) in A and reduced set (marked with red circles) at home ; 4-channel electro-oculography (EOG) in A and 2-channel in B; 1-channel electrocardiography (ECG) in A and 3-channel in B; Electrodermal activity (EDA) was recorded in A; At home chin electromyography (EMG) was recorded as part of polysomnography (PSG). The placement of electrodes (M) for mandibula area EMG is shown in picture with MZ as reference.

Patients had a short break between cognitive tasks to prevent fatigue from static sitting in front of a computer screen. Signal gathering was interrupted during breaks and when changing from one task to another as movements of subjects caused artefacts to the signals. Gathering of physiologic signals was restarted a few minutes before each task session.

To determine individual baselines for the different physiologic measurements, we first gathered data while subjects watched a non-stressful nature video for 7 minutes. After the 80-minute study session, this 7-minute session was repeated. Then using ERP recordings, a 30-minute passive multi-feature mismatch negativity (MMN) recording was carried out to study central auditory processing, namely change detection and involuntary attention allocation to emotionally valenced speech stimuli [12].

Before starting the laboratory day, the subjects rated their subjective level of sleepiness with the Karolinska Sleepiness Scale (KSS) [13] and between the different computerized tasks, six times in total. Participants also estimated (4 times in total) their own test-related performance and perceived task load with the NASA Task Load Index (NASA-TLX) [14]. More details on laboratory day methods are presented in the online <u>supplemental 2</u>.

#### Home measurements

The home measurement protocol is presented in figure 4. The 1^st^ and 3^rd^ were carried out directly after the laboratory tests and the 2^nd^ in between. FIOH research nurses attached the sensors recording different aspects of physiology also for the 2^nd^ recording taking place only at home. The patients performed the portable, tablet-based Flexible Attention Test (FAT battery) in the evening and next morning. This computerized test set is designed to measure several cognitive functions, including visuomotor speed, attention, executive functions, and working memory. FAT has been developed by Dr Paajanen at FIOH. It includes eight relatively short subtests evolved from classical neuropsychological tests: Trail making test and Corsi Block Tapping task [6]. The other tablet-based cognitive test was the Verbal Paired Associates test [15] designed to measure verbal memory, learning and over-night memory consolidation. Participants performed learning and immediate recall parts of the test in the evening and the recall phase again the next morning after night sleep. The next day the patients re-visited FIOH to return the devices and give their feedback on the study.

**Figure 4.**
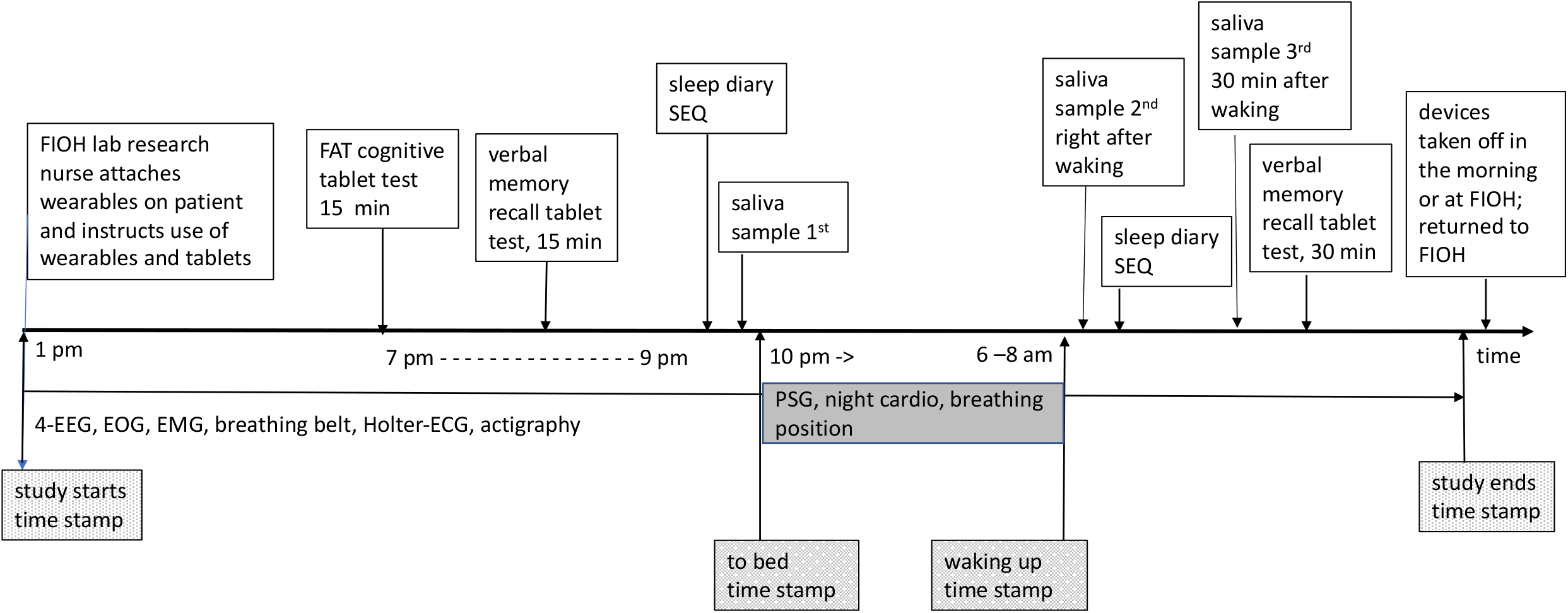
Study protocol of home recordings. Home recordings protocol. FIOH = Finnish Institute of Occupational Health, FAT = cognitive battery and Verbal memory test, EEG = electroencephalography, EOG = electro-oculography, EMG = electromyography, ECG = electrocardiography, PSG = polysomnography, SEQ = self-energy-questionnaire.

The attachment of physiological monitoring devices is shown in figure 3 b. The setup was chosen especially for doing sleep polysomnography (PSG) at home. With this reduced set of EEG channels, EOG and chin EMG recordings sleep can be classified into different stages [16]. In addition, a whole night 3-channel ECG-Holter recording was included in the setup as heart rate and heart rate variation (HRV) have been reported to provide information on the level of recovery during sleep [17]. Heart arrhythmia indicative of heart disease often occurs during early morning sleep [18].

In the evening, patients reported their estimated day workload level in the sleep diary and in the morning added information on perceived quality and quantity of sleep. They answered the Self-Energy-Questionnaire (SEQ) [19] in the evening and after waking up, and 30 minutes later. Patients collected three cortisol saliva samples, one near bedtime and two in the morning, 30 minutes apart.

Actigraphy combined with a sleep diary is often a first stage screening procedure in patients with sleep problems [20]. Thus, these were included in each home recording protocol. The 4^th^ home measurement day, however, only consisted of a sleep diary and actigraphy which were posted to the patients from FIOH. Patients sent them back to FIOH together with the recordings.

#### Health questionnaires

Patients filled in questionnaires linked to the laboratory and home measurement studies. Patients were also asked to fill in repeatedly validated digital versions of subjective questionnaires via an internet portal. They covered health habits, perceived mental health, fatigue, burnout, stress, sleep quality, subjective cognitive symptoms, cognitive performance, physical activity and everyday performance at home and work. The Brain Work Questionnaire [21], recently developed at FIOH focusing on frequency of cognitive demands and appraised strain at work was also included in the set. Figure 1 shows the planned time points when patients were sent phone messages with requests to provide answers. Detailed list of questionnaires and references is provided in the online <u>supplemental 3</u>.

#### Assessment of patients’ study experiences

Research nurses gathered feedback on patients’ subjective experiences concerning the study. Patients also filled in a questionnaire after each examination day on protocol-related mental and physical strain, required effort and possible experienced frustration, opinions on the number of different tests and measurements, as well as the clarity of instructions. Details are presented in the online <u>supplemental 3</u>.

#### Data management

Data were collected using a registry designed for this study by BCB Medical. Details are described in the online <u>supplemental 2</u>.

Though the study was started before the new General Data Protection Regulation Data (GDPR) guidelines came into force, we addressed patient privacy issues accordingly. Concerning this study, patient identification data (name and social security number) were stored in a separate data repository which also included the number code given to the patient. All outpatient laboratory and home recording data were anonymized with an algorithm before they were uploaded into a research data repository of HUS. As a mandatory safety measure, the number code was attached to a patient’s measurement data. Thus, only the treating physician can use the code to identify the patient from the separate privacy repository if further analysis of research data detects a possible medical problem needing further examinations. This later access to the patient’s data is marked into the repository’s logbook.

The signals were recorded using multiple devices resulting in data files in multiple formats. The data files were combined into one data file in the HDF5 format using a software developed in this project for this purpose (the export2hdf5 programme, https://github.com/bwrc/export2hdf5). The resulting fused physiologic data in HDF5 format was uploaded to the data repository.

Clinical neuropsychological data was not uploaded into the repository as analysis will be carried out separately by trained clinical neuropsychologists. Data with medical information of the patients were in separate sub-repositories of the research repository. Without knowing the identity of the patients, researchers working in different disciplines can later analyse data stored in the HUS data repository with a wide variety of data science methods. Figure 5 shows the data repository development and structure.

**Figure 5.**
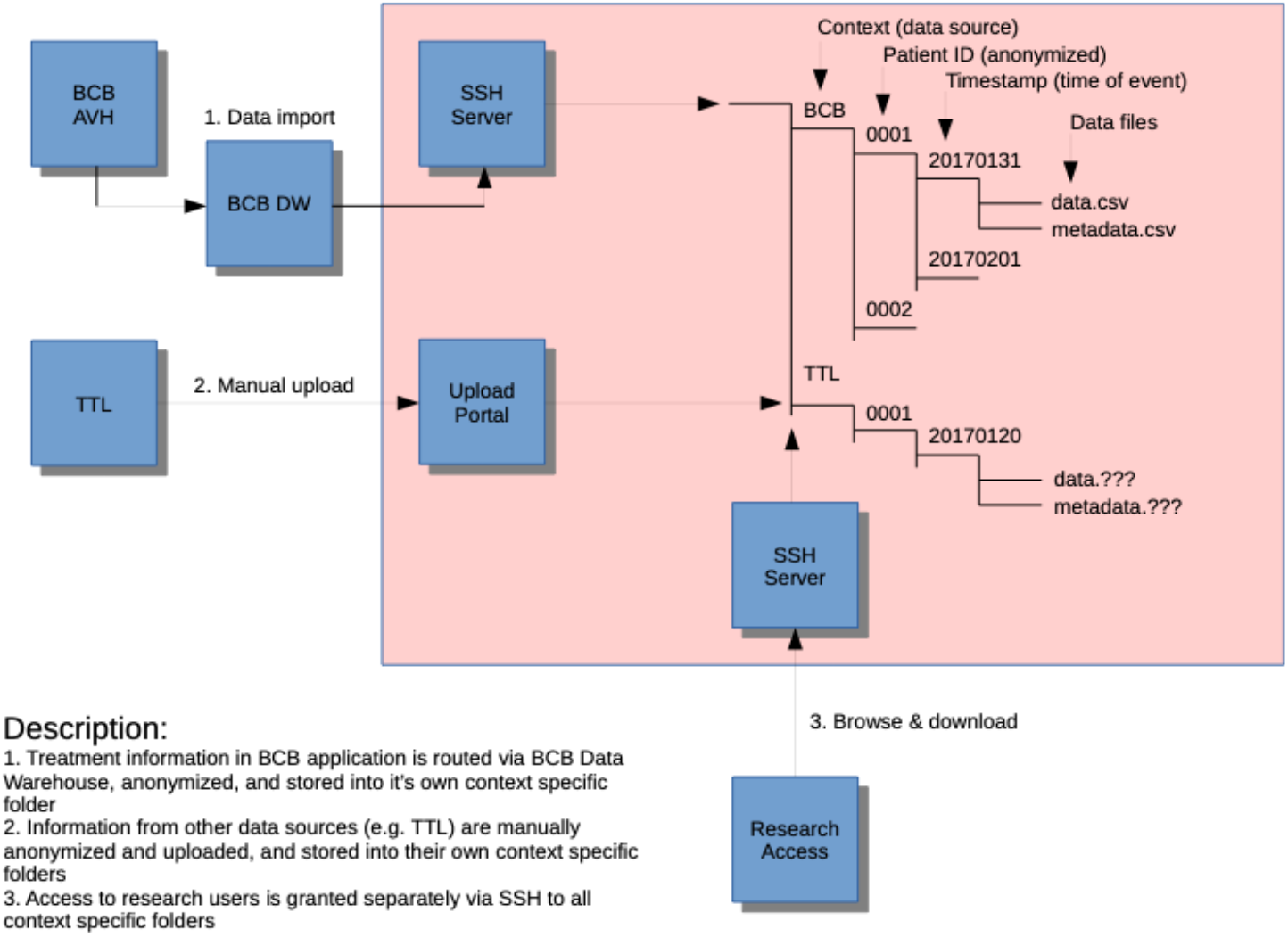
Data repository developed for the study by BCB Medical. Data repository developed for the study by BCB Medical.

### Executability and repeatability measures

The following outcome measures were used to evaluate the success of carrying out the study protocol: 1) Number of patients completing the whole or part of the study protocol, 2) Actual study time points in relation to those planned in the protocol, 3) The success rate of obtaining data with the different examination methods, 4) The amount of physiologic data gathered from the patients during home recordings. In addition, qualitative information on patients’ study experiences were gathered.

## RESULTS

### Number of patients completing the study protocol

All 16 patients participated in pre- and postoperative clinical neuropsychological tests and the preoperative laboratory and home recordings. In one patient only a preoperative actigraphy was done at home. This patient did not continue in the study after the UIA operation. Another patient could not continue because of a postoperative stroke. Thus 14 (87%) patients participated also in the 1^st^ postoperative home recordings and 13 in the 2^nd^ postoperative day examinations, but 2 of them only in the home recording part. Thus, all planned home recordings (including PSG, ECG, and cognitive tablet tests) were carried out in 13 (81%) patients. Eleven (67%) patients participated in all 4 examination days as planned. All obtained basic data was determined to be of good quality (visual inspection). For more details see table 1 and its footnote.

**Table 1.**
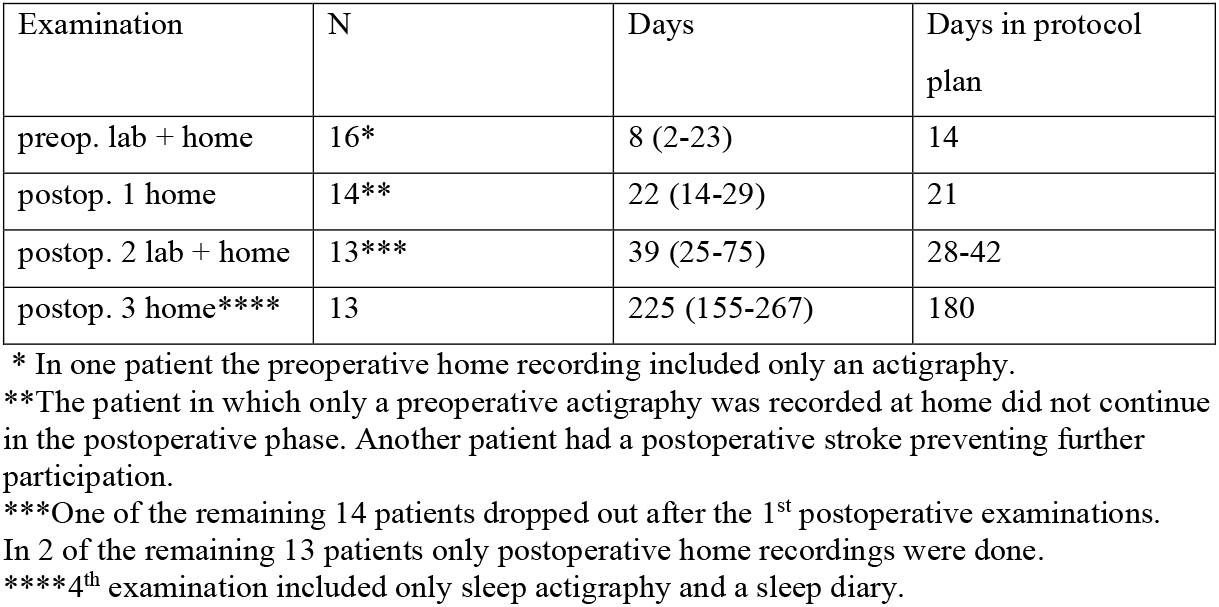
Actual data gathering time points (expressed in days, median and range) in relation to the operation day (before = preop. and after = postop.) and the original planned time points for the examinations. N = number of patients, lab = laboratory.

Table 1 also shows the actual time points of data gathering. They differed from the preplanned pre- and postoperative times (in days) shown in Figure 1.

### Success rate of gathering planned data sets

All planned data was gathered successfully and in a similar way during the 2 laboratory days from those subjects examined.

A total of 13 patients participated in all 4 home recordings. In one patient the preoperative home recording consisted only of the actigraphy and in one the preoperative ECG home data was missing. In all other subjects all data was obtained as planned. Thus, we achieved 41 successful home PSG and 40 Holter ECG recordings with the same sensor/device setup. All tablet cognitive test data was obtained as planned from all 41 patients examined at home. Details are provided in table 2.

**Table 2.** Total amount of physiologic data in minutes acquired with home recordings.

| Physiologic metric | N | median | range |
| --- | --- | --- | --- |
| EEG | 41* | 1070 | 786-1540 |
| ECG, Holter | 40** | 1115 | 837-1374 |
| PSG | 41* | 490 | 365-630 |
| Actigraphy *** | 40 | 1347 | 1085-1617 |
| ****Actigraphy including 4<sup>th</sup> exam | 53 | 483 | 345-680 |

\*In 13 patients all three home recordings with also polysomnography (PSG) were done. In one patient only the preop and 1<sup>st</sup> postoperative examination were done. Thus, the total N is 39 + 2.
\*\*In 1 patient preoperative electrocardiogram (ECG) was missing.
\*\*\*Actigraphy included in the setup with electroencephalography (EEG), ECG and PSG, was missing in 1 patient in the 2<sup>nd</sup> postoperative recording
\*\*\*\*Actigraphy in all 4 recordings, 4<sup>th</sup> one missing in 1 patient
The placements of sensors of the different recordings are shown in Figure 3.

Patients filled in all questionnaires and diaries implemented into the examination days. They included NASA-TLX and KSS during laboratory examinations and during home examinations the sleep diary and SEQ questionnaire. Only 30 % of the patients filled in all the questionnaires in the internet portal.

### Patients’ feedback on the study

Most patients did not experience discomfort from using the different wearables. Moreover, EEG sensors near the neurosurgical wound site were tolerated well. During home recordings, the patients experienced moderate, a few notable, difficulties in performing on their own the cognitive tablet tests. Patients did not find salivary sampling difficult or cumbersome. Most patients reported at least moderate mental and physical workload, frustration, and increased effort after both laboratory days. The postoperative laboratory day was reported to cause slightly more physical and mental workload than the preoperative one.

Concerning the laboratory and home recording protocols, patients evaluated the number of tests and questionnaires good, and instructions clear for all examination days. The questionnaires to be filled via the internet portal were considered too extensive and answering time too long.

The few patients who did not complete the study felt that instructions were insufficient, possibility to ask help was inadequate and the number of tests was too high. See online <u>supplement 3</u> for more details.

## DISCUSSION

We examined the executability and repeatability of this multiparametric study protocol in patients undergoing UIA neurosurgery. The group was chosen as neurosurgery is a medical intervention with a clear time point. Thus, accurate times of the pre- and postoperative examinations could be defined. Also, recovery from operation differs between UIA patients. Our aim was to test the use of different wearable and mobile technologies to gain experience on their usability in obtaining objective data on different aspects of performance during follow-up of recovery.

Undergoing brain surgery for UIA is mentally and physically demanding for the patient. Despite this, our results show that many patients were able to go through the whole challenging measurement protocol. Fourteen (87%) out of 16 recruited patients participated in pre- and postoperative examinations, 11 completing the whole protocol and 3 partly. Nearly all planned data was obtained during the out-patient laboratory test sessions. The research nurses’ role was crucial for this success.

Missing data (Tables 1 and 2) from home recordings was mostly due to a totally missing home recording day. The role of research nurses attaching the wearables on the patients and giving detailed instructions was important. During home recordings, patients could contact research nurses if needed. This working mode presents a significant workload on nurses. For wider use of medical grade technologies outside hospitals and special laboratories, we need easy-to-use solutions. To obtain good signal quality patients themselves need to be able to place the sensors accurately on the planned body areas and to check that the device is functioning properly.

In our study each patient acted as their own control. We chose a setup in which patients are tested before and after a medical intervention. Without baseline knowledge of a patient’s performance data prior to an intervention, it is difficult to deduce what observed changes might be due to treatment and recovery. Baseline performances varies between individuals and thus the use of performance values based on group studies can be unreliable.

The physiological methods implemented into the study setup were chosen based on earlier research on their ability to reliably capture changes in human physiology. EEG is a widely used technique to dynamically evaluate brain function during sleep [22], sleep deprivation and vigilance [23], attention, and cognitive load [24,25]. Eye movements also capture daytime fatigue and sleepiness [26,27]. EEG activity increases in response to increase in work task demands [24].

Eye movements and attentional functions share common neural networks in brain frontal areas [28]. Therefore, eye movements have been used to study attentional functions, as well as vigilance [10,29]. Electrodermal activity (EDA) reflects activity of the sympathetic branch of the autonomic nervous system i.e., arousal and an acute stress response [30], and is a widely used metric for estimating stress [31]. Mobile ECG for calculating heart rate variability is used to evaluate the effects of different types of stress on the autonomic nervous system [32]. Heart rate variability can be used to evaluate an individual’s stress response during a cognitive task [33]. Enhanced sympathetic nerve activity due to a decrease in parasympathetic nerve activity is common in acute, sub-acute, and chronic fatigue [34].

Sleep disturbances are common [35,36] and cause daytime fatigue and weariness. Deep sleep is especially important for recovery [17]. Night-time heart rate variation is a good indicator of cardiovascular recovery during sleep [37]. PSG is needed for accurate sleep stage classification and was thus included in our home recording setup. As cardiovascular problems are often captured during night [38,39], we recorded the whole night ECG synchronizing it with PSG. The reproducibility of PSG and ECG was good; in only one patient the home ECG was missing.

The combination sleep diary and an actigraphy measuring movements is used as a first stage clinical screening method in sleeping problems [40]. With our study setup it is possible to study the relationship between these screening methods and findings in PSG and whole night ECG.

Daytime mobile ECG can be used as biofeedback during recovery from operations and illnesses to fine-tune activities according to a person’s available resources [41]. Cardiovascular diseases can present themselves as excessive fatigue and inability to allocate more physiologic resources to more demanding tasks. The wide range of both physiologic and medical conditions affecting HRV needs to be kept in mind [42].

Saliva cortisol levels are used as a stress biomarker even though the hormone levels are affected by a wide range of internal and external factors [43,44]. With our study setup relationships between different physiologic variables and cortisol levels can be studied.

Questionnaires and diaries linked to mobile device recordings worked best in this study. Questionnaires presented via an internet portal followed a separate timeline and only 30 % of patients answered them fully. Identifying questionnaires with a best fit-for-purpose could result in a better answering percentage. The lesson here is that the questionnaires not linked to actual measurements should be piloted beforehand which we did not do.

An important outcome of this study is the data repository. It contains all raw data with time stamps and annotations, (currently) further processed ECG and PSG data, computerized task performance outcome measures and subjective questionnaires filled in at the time of medical device-based measurements. The data can be further examined with different clinical and data science approaches. A system confirming correctness of the data uploaded from different device data loggers was implemented. The data labeling protocol not only considers the type of data, but also at what time point the data was gathered in the project (pre-or postoperative, laboratory or home recording).

More studies on patient self-monitoring can identify usability issues that, if not considered, decrease the quality of data. Remote monitoring of device performance is needed to identify and prevent erroneous use. In addition, while ensuring patient privacy is highly important, it is equally important to make sure that the device is used in a correct manner by the patient it has been allocated to. Secure data streaming requires a data hacking and tampering identification solution [45].

## CONCLUSIONS

This study shows that a wide variety of both physiological and cognitive performance data can be obtained from a patient before and after a medical intervention and during follow-ups, also outside the hospital. The UIA patients studied were a selected volunteer group. More comprehensive experience on usability and patient compliance is needed by examining other patient groups Accumulating data from a patient and different patient groups can be used 1) to identify the best combination of outcome performance measures that provide information on how a person performs in everyday life, 2) to understand to what extent measurement results vary between individuals and 3) to determine what is the best, most informative combination of performance measures in an individual for studying and predicting efficacy of medical treatments, forms of rehabilitation, disease progression and recovery, as well as working ability. This knowledge also paves the way to personalized medicine.

## Supporting information

Patient recruitment flow chart

Psychological methods

Method descriptions

## Data Availability

The data gathered during this research is anonymised and stored in a data repository at Helsinki University Central Hospital. This paper describes only outcome data relevant for evaluating the executability and reproducibility of the study protocol.

## Contributors

Conception of the study (KM, MK, JP, SP), study design and methods (KM, SP, JP, KP, LS, JV, LA, HJ, TP, AA, MK, NM-M), data analysis (KM, SP, KP, LS, TP, JV, JK, AH, IR), data repository and data storage solutions (AH, JK, TT, AM, KM, JP, MK), patient clinical examinations (AA, MK, NM-M). KM is responsible for writing the article. IR participated in writing the first draft. All named authors participated in revising the manuscript, and all have approved the final version to be published.

## Acknowledgements

We thank M.Sc. Mika Tervonen for providing data repository expertise, psychologist Michaela Salo for participating in neuropsychological assessments of subjects and research nurses Laura-Leena Kupari (HUS), Nina Lapveteläinen (FIOH), and Riitta Velin (FIOH) for their invaluable and skillful work in ensuring successful practical carrying out of this study.

