## Supplementary material for "Executability and repeatability of a study setup using wearable and computerized technology to examine a wide range of physiological and cognitive functions of a patient outside hospital": Patient recruitment flow chart

Kiti Müller et al.

### SUPPLEMENT 1

*Flow chart of the patient selection process, outcome of recruitment and patient participation in the actual study.*

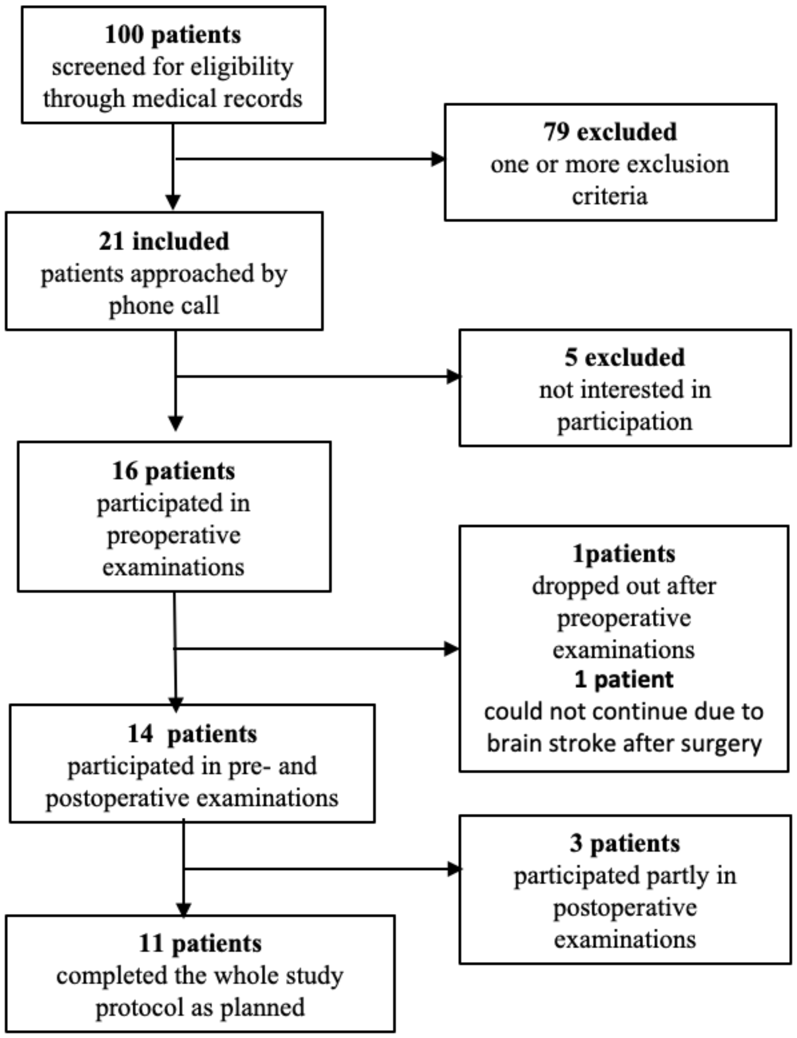
