## Supplementary material for "Executability and repeatability of a study setup using wearable and computerized technology to examine a wide range of physiological and cognitive functions of a patient outside hospital": Psychological methods

Kiti Müller et al.

**SUPPLEMENT 3**: **Clinical neuropsychological tests and subjective questionnaires**

Table 1. Clinical neuropsychological test battery

| **Cognitive domain** | **Neuropsychological test** |
|---|---|
| Processing speed | WAIS-IV Coding [1]<br>Stroop test, colour-congruent [2]<br>FAT, perceptual motor speed [3,4] |
| Executive functions | Stroop test, colour-incongruent [2]<br>Dual task [5]<br>Verbal fluency test, phonemic [2]<br>FAT, set shifting [3,4] |
| Working memory | WMS-III Letter-Number Sequencing [6]<br>WMS-III Digit Span [6]<br>FAT, visuospatial span [4] |
| Memory and learning | WMS-III Word Lists [6]<br>WMS-III Logical Memory [6]<br>WMS-III Visual Reproduction [6] |
| Visuospatial perception | WAIS-IV Block Design [1]<br>Visual search for parallel lines [7] |
| Verbal functions | WAIS-IV Similarities [1]<br>Boston naming test [8]<br>Verbal fluency test, semantic [2] |
| Questionnaires | Checklist for cognitive and emotional consequences following stroke (CLCE-24) [9]<br>Cognitive Reserve Scale (CRS) [10]<br>Resilience Scale (RS-14) [11] |

Table 2. Subjective Questionnaires

Patients answered the following validated subjective questionnaires via an internet portal.

| **Questionnaires and references** | **Short description** |
|---|---|
| Oslo Social Support Scale [12] | Three-item scale used to determine the social support of individual's family, friends, and neighbours |
| Alcohol Use Disorders Identification Test (AUDIT) [13] | Widely used 10-item screening tool developed to evaluate individual's alcohol-related problems |
| WHO Alcohol, Smoking and Substance Involvement Screening Test (ASSIST) [13] | A brief screening tool used to discover the use of psychoactive substances |
| International Physical Activity Questionnaire (IPAQ) [14] | A brief questionnaire for assessing individual's health-related physical activity comprehensively and multidimensionally |
| Mediterranean Diet Score (MDS) [15] | Nine-item score to evaluate the healthiness of individual's diet habits |
| Shirom-Melamed Burnout Measure (SMBM) [16] | Fourteen-item tool to evaluate individual's burnout level |
| Absenteeism and Presenteeism questions of the World Health Organization's Health and Work Performance Questionnaire (HPQ) [17] | Seven-item questionnaire extracted from WHO's HPQ. Describes individual's work performance and the number of sick leaves and other work absences |
| Perceived Stress Scale [18] | Ten-item scale to describe stress experiences |
| Mobile Modified Rankin Scale [19] | Mobile-based scale used to evaluate performance and independency mostly in stroke patients |
| Beck Depression Inventory (BDI) [20] | Widely used questionnaire to evaluate the level of depression symptoms |
| Beck Anxiety Inventory (BAI) [21] | Widely used questionnaire to evaluate the level of anxiety symptoms |
| Epworth Sleepiness Scale (ESS) [22] | Ten-item scale describing day-time sleepiness |
| Basic Nordic Sleep Questionnaire (BNSQ) [23] | A questionnaire determining a wide range of sleep complaints |
| Fatigue Assessment Scale (FAS) [24] | Ten-item scale evaluating the levels of cognitive performance and fatigue |
| EuroHis-8 [25] | Eight-item questionnaire to evaluate an individual's quality of life |
| Checklist for Cognitive and Emotional consequences (CLCE-24) [9] | A screening tool to identify cognitive and emotional problems |
| Brain Work Questionnaire (BWQ) [26] | A comprehensive questionnaire evaluating cognitive demands at work and work-related cognitive strain |

Table 3. Evaluation form filled after home recordings. Estimates are median values (0-10) with interquartile range (IQR). The items of “Number of tests” and “Number of questionnaires” are graded from 0 = too many to 10 = too few and 5 represents not too many or too few. Otherwise, the higher the value the easier or more positive experience. (N= number of patients answering.)

| **Evaluation about the study protocol** | **Preoperatively (N=8)** | **1st postoperatively (N=11)** | **2nd postoperatively (N=11)** |
|---|---|---|---|
| Mental demand | 7.5 (7-10) | 10 (4-10) | 6 (4-8) |
| Physical demand | 8.5 (7-10) | 9 (6-10) | 6 (5-9) |
| Effort | 7 (4-8.5) | 9 (4-10) | 6 (5-8) |
| Frustration | 8.5 (7-9.5) | 10 (4-10) | 5 (4-10) |
| Saliva samples | 10 (9-10) | 10 (10-10) | 10 (5-10) |
| Sleep diary | 10 (8.5) | 10 (10-10) | 10 (8-10) |
| Cognitive tablet tests | 9 (5-10) | 10 (8-10) | 9.5 (9-10) |
| Physiological measurements | 9.5 (9-10) | 10 (10-10) | 10 (8-10) |
| Instructions<br>● Sufficiency<br>● Clearness | <br>10 (9.5-10)<br>10 (10-10) | <br>10 (9-10)<br>10 (9-10) | <br>10 (10-10)<br>10 (9-10) |
| Opportunity to ask help | 10 (8-10) | 10 (10-10) | 10 (10-10) |
| Number of tests | 5 (4-5) | 5 (5-5) | 5 (3-5) |
| Number of questionnaires | 4.5 (3.5-5) | 5 (4-5) | 5 (2-5) |
| Re-participation | 10 (8-10) | 10 (6-10) | 10 (5-10) |
