## Supplementary material for "Executability and repeatability of a study setup using wearable and computerized technology to examine a wide range of physiological and cognitive functions of a patient outside hospital": Method descriptions

Kiti Müller et al.

**SUPPLEMENT 2: Outpatient laboratory examination methods and data management**

### Physiological measurements

#### Amplifier specifications

- NeurOne amplifier (Bittium, Oulu, Finland)
- Sampling rate 500Hz, lowpass digital filtering 0-125Hz, for all channels
- AC mode: Analog high pass filter (-3dB point): 0.16Hz
- EEG, EOG, ECG, respiration: AC measurement (0.16Hz-125Hz)
- EDA: DC mode (0–125 Hz)
- Common reference and ground were placed at electrode sites FCz and AFz

#### Electroencephalography (EEG)

The EEG was recorded continuously using a 32-channel active electrode system (actiCAP, Brain. Products GmbH, Gilching, Germany) connected to a NeurOne amplifier. EEG data was collected from 28 electrodes (Fp1, Fp2, Fz, F3, F4, F7, F8, FC1, FC2, FC5, FC6, Cz, C3, C4, T7, T8, TP9, CP1, CP2, CP5, CP6, TP10, Pz, P3, P4, P7, P8, Oz) placed according to the international 10–20 electrode system (see Figure 1). The common reference and ground were placed in the electrode cap at electrode sites Fcz and Afz, respectively. Two electrodes were placed at the left and right mastoids to allow referencing in later analyses.

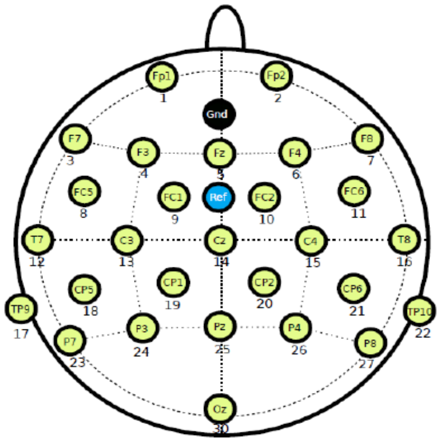


Figure 1. Electrode placements (yellow): Fp1, Fp2, Fz, F3, F4, F7, F8, FC1, FC2, FC5, FC6, Cz, C3, C4, T7, T8, TP9, CP1, CP2, CP5, CP6, TP10, Pz, P3, P4, P7, P8, Oz, ref (blue): common reference (Fcz) and Gnd (black): ground Afz.

Electro-oculography (EOG)

Horizontal and vertical EOG were measured with four Ag-AgCl electrodes placed at the outer canthi of both eyes (right: heogr, left: heogl), as well as above (veogu) and below (veogd) the left eye (see Figure 2). The measurement was monopolar, and the electrodes were referenced to the common reference.

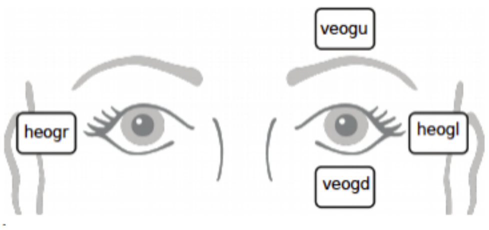


Figure 2. EOG electrode placement.

Electrocardiography (ECG)

Electrocardiography (ECG) was measured with two electrodes from the chest (bipolar coupling), one placed on right side of the body under the collarbone and another electrode on the left side of the body on the rib cage (see Figure 3).

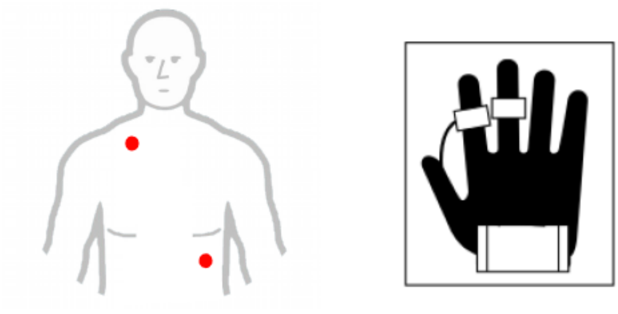

Figure 3. (Left) ECG electrode placement, (right) EDA electrode placement.

Electrodermal activity EDA

Electrodermal activity (EDA) was measured using mt-gsr-1 sensor by Mega Electronics. Two dry Ag-AgCl electrodes were placed on right index and middle fingers, facing the volar surface on the medial phalange (Figure 3). The sensor operates with one channel and conductance between sensors was collected using Neurone system in DC mode.

Respiration

Respiration was measured using two respiration belts (Embla XactTrace Reusable Universal RIP Belt, Natus neurology, USA); one sensor placed around the abdomen (res_abdomen) and the second sensor around the chest (res_thorax).

### Cognitive tasks

Laboratory environment

The measurements were executed in a soundproofed measurement chamber. The participants were placed in a chair at a 74 cm distance from the computer screen. Sounds were presented via two loudspeakers (Genelec, Iisalmi, Finland) placed on the wall of the chamber at a height of 110 cm and at 115 cm from participants. The loudspeakers were placed at approximately 30 degrees to the left and right of the participant.

All stimuli were presented within the same stimulus sequence using the Presentation software (Neurobehavioral Systems Inc., version 17.2). The stimulus software sends port codes directly to the NeurOne's markers channel.

Baseline (BL1, BL2) measurements

In the baseline measurement participants were instructed to relax and watch a muted video film. Each baseline measurement lasted 6.5 minutes.

N-Back (NB0b, NB1b, NB2b)

N-back task [1] was carried out in this study as follows: It consisted of two types of stimuli: visual and auditory. Visual stimuli were bright white numbers (0-9) on a black background (static contrast ratio 1000:1), presented in the centre of a computer screen subtending a visual angle of 1 x 1.9 degrees at a distance of 74 cm in front of the participant. Duration of the visual stimulus was 500 ms, and the stimulus-onset asynchrony (the amount of time between the onset of one stimulus and the onset of the next stimulus) was 2000 ms. During the 1500 ms delay period a black screen was visible. The visual N-back task consisted of 0-, 1-, 2-back conditions. Each participant completed the conditions in the same, previously presented, order. In the 0-back condition participants indicated whether the presented stimulus was a predetermined target '5'. Participants were instructed to respond to the target stimuli by pressing a button with the right index finger, and to the non-target stimuli by pressing a button with the left index finger. In the 1-back condition, the participants task was to monitor whether the current number was the same (target), or not (non-target), as the previous one. In the 2-back condition, the current stimulus was compared with the stimulus presented two trials before. Each condition consisted of a total of 212 visual stimuli of which 33% were target and 67% were nontarget stimuli - presented in a random order.

Sounds

In parallel with visual n-back tasks, participants were presented with 96 ecologically valid naturalistic distractor sounds (e.g., produced by a hammer, drill, telephone ringing, door, rain etc). Each distractor sound was presented once across all the conditions (32 per condition). Sound duration was 200 ms with intensity of 57 decibels (dB) sound pressure level (SPL) on average [2,3].

Notes on stimuli

Here target is a stimulus at position i which is equal to:
NB0b) stimulus to detect (position i)
NB1b) preceding stimulus (position i-1)
NB2b) stimulus at position i-2

The stimulus pairs {i-1, i} and {i-2, i} form a pair hereafter called the "detection pair".
To compute an ERP for the stimulus that _ends_ the detection pair, the epochs are classified using the immediately following response. In other words, presentation keeps track of what is the correct answer for each of the stimuli. To compute an ERP for the stimulus that _starts_ the detection pair, the epoch classification needs to take the time lag into account.

Instructions

Prior to each condition, participants were presented with written instructions and a practice trial to familiarize themselves with the tasks. Between conditions, the participants had a brief break. Task performance (i.e., accuracy, reaction times) metrics were recorded. Participants did not receive any feedback on their performance.

Task switching (TS1, TS2, TS3)

The task switching tests [4] were carried out as earlier described [5]. The stimuli were white letter-number pairs on a black background subtending a visual angle of 2.4 Å~1.6Åã. The stimulus pairs consisted of a defined subset of Arabic numbers (2–9) and Latin letters (A, E, I, U, G, K, M, R). A solid, white horizontal stationary line (height 0.3Åã, length 5.6Åã) was present at the centre of the screen. Each letter-number pair was presented either above or below the line in pseudorandom order with the letter always presented on the left side of the pair. The position of the letter-number pair was always either above or below the horizontal line (with a vertical gap of 0.5Åã between the letter-number pair and the line), and this served as a cue according to which the participants were required to judge the stimulus pairs: when the stimulus pair occurred above the horizontal line, the participant had to decide whether the number in the letter-number pair was even or odd. When it occurred below the line, the participant had to classify the letter as vowel or consonant.

The decision to be made in each task was unknown to the participant until the letter-number pair was presented. The participants were instructed to respond to each stimulus pair with a keyboard button press (left and right ctrl buttons covered with tape were used): vowels and even numbers required a response with the right index finger, consonants and odd numbers with the left index finger. Speed and accuracy of response were equally emphasized in the task instructions. The pairing of the letter-number combinations (e.g., 'E5') was semi-randomized so that approximately half of the character pairs were incongruent and half of them were congruent, that is, the task-irrelevant character was mapped either to a response with same or the other hand (e.g., when the task was to classify the number as odd or even in 'E5', a correct response was given with the left hand, whereas the task-irrelevant character was mapped to a correct response with right hand).

The task consists of three phases:

1) Reaction time (TS01): Participants were instructed to push the keyboard button immediately when the letter-number pair appears on the screen
2) Recognition (TS02): a) Letter-number pairs were presented above the horizontal line and participant's task was to recognize if the number was even or odd. b) Letter-number pairs were presented below the horizontal line and participant's task was to recognize if the letter was vowel or consonant. Conditions were not mixed at this phase.
3) Task switching (TS03): Participants were required to judge the stimulus pairs according to the position (above or below horizontal line) of the letter-number pair.
In total, the paradigm consisted of 545 stimulus pairs with 122 task switches (22%) and 423 task repetitions (78%). Task runs of one to nine stimulus pairs were presented in succession above or below the line before a switch. In the entire sequence, there were 20 task runs (16%) consisting of only one stimulus pair before a switch, 26 task runs (21%) constituting two repetitions before the switch, and on average 11 task runs (9%) of 3 to 9 repetitions, each. Each stimulus pair was shown

until response, however, not longer than 2500 ms. The presentation rate was tied to the participant's response in the following way: a correct response was followed by a 150 ms delay period after which the next stimulus pair was presented. An incorrect or missed response was followed by a 1500 ms delay until the next stimulus pair was presented.

### Saccade task

The setup has earlier been used in the study developing saccade analytics for identifying fatigue [6]. Participants were instructed to sit still, avoid blinks, and to look at the location of the central fixation point until the target stimulus appeared, after which they were supposed to move their gaze as quickly as possible to the target stimulus. When the stimulus disappeared, they were instructed to move their gaze back to the central fixation point. The saccade was 10 degrees of visual angle whereas the size of the fixation point and the target stimulus were 1 degree of visual angle. The saccade task consisted of alternating A (overlap stimulus) and B (gap stimulus) blocks. In the A block the central fixation point was visible all the time.

The task sequence started with the central fixation point. After 1000 ms the target stimulus appeared on either the left or the right side of the screen. The target stimulus disappeared after 1000 ms and the next trial started with a central fixation point. In the B block the fixation point disappeared 200 ms before the target stimulus appeared. The saccades were presented in 20-saccade blocks and there were 5 s rest pauses between the blocks. One 20-saccade block lasted 40 s in the A block and 44 s in the B block. In each 8-minute measurement session 200-saccade stimuli, 100 per condition (A or B) were presented. Here, measurements start with the B block (stimulus sequence: B_A_B_A_B_A_B_A_B_A).

### Multi-feature mismatch negativity (MMN)

The participants conducted the multi-feature MMN paradigm[7,8] with standard, 9 deviant and 3 novel stimuli with emotional prosody. The presented stimuli were as follows: The standard stimulus was a 336-ms natural utterance of a bisyllabic Finnish pseudoword /ta-ta/, with the stress on the first syllable as indicated by a slightly higher F0 (fundamental frequency, i.e., pitch) and intensity compared to the second syllable. The deviants differed from the standard in spectral density, frequency, intensity, sound-source location (left/right), noise level, consonant duration (/ta-t:a/), omission (/ta-/), vowel change (/ta-to/), or the vowel duration (/ta-ta:/) of the second syllable of the pseudoword (referred to as Den, Fre, Int, Loc, Noi, ConDur, Om, VowCha, and VowDur, respectively).

The deviation always occurred in the second syllable of the pseudoword, except in the location deviant in which the deviation appeared in the beginning of the pseudoword. The vowel-change deviant (/ta-to/), and the vowel-duration deviant (/ta-ta:/) were recordings of natural utterances, therefore the physical characteristics of these stimuli differed from the standard slightly on the first syllable too. The other seven deviants were created by digitally editing the standard stimulus and were thus identical to the standard except for the edited auditory attribute. The three stimuli with strong emotional prosody i.e., happy, angry, and sad were used as rarely occurring novelty-like variants of the standard. Physical characteristics of the stimuli differed from the standard considerably, for example in length, pitch, and momentary intensity.

The standard and deviant stimuli were presented 210 times each (P = 0.09, each), and the three emotional stimuli were presented 42 times each (P = 0.02). The stimuli were pseudo-randomized so that neither the same deviant type nor the standard was repeated consecutively, and the emotional rare sounds were presented in varying intervals, once every 10-16 seconds. The stimulus onset asynchrony (SOA; the amount of time between the onset of one stimulus and the onset of the next stimulus) was 750 ms, and the total recording time 28 min.

During the recording, the participants watched a muted video film. They were instructed to relax, avoid excessive and unnecessary blinking, muscle tension and movements, as well as to ignore the sound stimuli.

### Subjective questionnaires

#### Karolinska sleepiness scale (KSS)

Patients filled in the Karolinska Sleepiness Scale (KSS) [9] before and after each cognitive task. KSS is a nine-point sleepiness scale, which estimates the psychophysical state experienced by the person in the last 10 minutes.

Scale description:

1) Extremely alert
2) Very alert
3) Alert
4) Rather alert
5) Neither alert nor sleepy
6) Some signs of sleepiness
7) Sleepy, but no difficulty to stay awake
8) Sleepy, some effort to keep alert
9) Very sleepy, great effort to stay awake or fighting sleep

NASA Task Load Index (NASA-TLX)

Patients filled in the computerized NASA Task Load Index questionnaire (NASA-TLX) [10] after the cognitive task sessions. This subjective, multidimensional assessment tool rates perceived workload on performing a certain task. The total workload is divided into six subscales:
1) Mental demand: How much mental and perceptual activity was required? Was the task easy or demanding, simple or complex?
2) Physical demand: How much physical activity was required? Was the task easy or demanding, slack or strenuous?
3) Temporal demand: How much time pressure did you feel due to the pace at which the tasks or task elements occurred? Was the pace slow or rapid?
4) Overall Performance: How successful were you in performing the task? How satisfied were you with your performance?
5) Frustration Level: How irritated, stressed, and annoyed versus content, relaxed, and complacent did you feel during the task?
6) Effort: How hard did you have to work (mentally and physically) to accomplish your level of performance?

The participants rated each task using a visual scale with a 100-points range with 5-point steps.

**Data management**

A neurologist worked together with a product manager and software provider of BCB Medical Ltd. First the stages of the data repository development project were addressed, including the used methods and schedule. The development process was carried out by using agile software development methods. In the specifications meetings, a functional prototype (mock-up) was built to make sure that the content and location of the data fields are appropriate in the upcoming registry.

The registries implemented by BCB Medical are integrated to hospital's information systems. The software provider in the project devised technical documentation of the registry specification. The documentation was used to execute the product development process of the registry. Before the final software launch the project team assessed the implementation solutions which provided valuable feedback for further development and enabled timely action for solving possible defects.

The first step in the deployment of the registry in Helsinki University Hospital (HUS) and other stakeholders was to configure the registry in the hospital's system. The system was tested and the registry's integrations to other medical data systems were launched. After this, the software provider organized training for the end users of the registry to ensure fluent deployment.

In adherence to General Data Protection Regulation Data GDPR each patient had at study enrolment been given an identification code that was used in all examinations to allow later reliable combining of a patient's data stored in different data set files in the repository.
